# The Impact of Education Level on the Risk of Heart Failure, Acute Myocardial Infarction, and Stroke in Patients with Atrial Fibrillation – a Swedish Nationwide Cohort Study

**DOI:** 10.64898/2026.04.05.26349791

**Authors:** Áron Sztaniszláv, Anna Björkenheim, Anders Magnuson, Nils Edvardsson, Dritan Poci

## Abstract

**Background:** Socioeconomic factors impact cardiovascular health. We investigated the association between patient education level and incident heart failure (HF), acute myocardial infarction (AMI), and stroke following a first hospitalization with atrial fibrillation (AF).

**Methods:** In this nationwide retrospective cohort study using linked Swedish national registers, we included all patients receiving a diagnosis of AF while hospitalized in Sweden from 1995 through 2008; categorized education level as primary, secondary, or academic; and followed patients for up to five years. Outcomes were first hospitalization for HF, AMI, or stroke. Associations were assessed using sex-stratified Cox proportional hazards models adjusted for age, calendar year of AF diagnosis, and measures of comorbidity burden (Charlson Comorbidity Index) and thromboembolic risk (CHA=DS=-VA score).

**Results:** The cohort comprised 263,172 patients (mean age 72.5±10.4 years; 56.2% male). Compared with primary education, secondary and academic education attainment were associated with lower adjusted risk of HF and AMI in both females and males. For HF, adjusted hazard ratios (HR) were 0.96 (95% CI 0.93–1.00) for secondary and 0.82 (95% CI 0.77–0.87) for academic education for females and 0.93 (95% CI 0.90–0.96) and 0.76 (95% CI 0.72–0.80), respectively, for males. For AMI, adjusted HRs were 0.89 (95% CI 0.85–0.93) and 0.71 (95% CI 0.65–0.78) for females and 0.91 (95% CI 0.87–0.94) and 0.75 (95% CI 0.71–0.80) for males. For stroke, lower adjusted risk was observed only in the academic education group. Baseline comorbidity burden and thromboembolic risk were higher in lower education groups.

**Conclusions:** Education level was inversely associated with risk of incident HF and AMI over five years, while the association with stroke risk was weaker. Documenting education level may help identify patients at increased risk who could benefit from careful monitoring and optimized preventive care.

## Introduction

Educational attainment impacts health literacy, cognitive abilities, and long-term income and is an important indicator of socioeconomic status (SES). Socioeconomic status, in turn, is a multifaceted factor in the incidence, prevalence, and outcome of cardiovascular diseases.[1,2,3] Atrial fibrillation (AF) is associated with substantial morbidity and mortality and imposes a significant economic burden, largely driven by hospitalization.[4,5] Although hospitalization rates for AF increased during the 1980s and 90s, more recent data suggest a declining trend in inpatient treatment.[6,7]

In a previous study, we found a significant inverse association between educational attainment and all-cause mortality among patients hospitalized with AF.[8] While mortality is a critical endpoint, morbidity outcomes may better reflect the clinical course following AF diagnosis. Morbidity data can also inform treatment efficacy and support clinical and policy decisions.[9]

Although associations between socioeconomic status, including educational attainment, and risks of heart failure (HF), acute myocardial infarction (AMI), and stroke are well described in the general population, evidence among patients with AF is limited.[1,10,11] This study aimed to investigate the association between education level reached and the risk of AF-related outcomes up to five years after an initial recorded diagnosis of AF during hospitalization.

## Methods

### Patient Population

This study was based on a previously described patient population.[12,13] We identified all individuals with a diagnosis of AF first recorded while hospitalized in Sweden from January 1, 1995, through December 31, 2008. Data were obtained by cross-linking the Swedish National Patient Registry and the Swedish Cause of Death Registry administered by the Swedish National Board of Health and Welfare, with the Total Population Register from Statistics Sweden.

Sweden has a publicly funded healthcare system and high-quality nationwide registries; the accuracy and validity of these registries have been evaluated previously.[14,15] Diagnoses were coded according to the International Classification of Diseases (ICD) using ICD-9 from 1987 to 1996 and ICD-10 beginning in 1997. The ICD codes for AF and study outcomes were defined as in previous studies.[12,13] We did not differentiate among paroxysmal AF, persistent AF, permanent AF, and atrial flutter.

### Study Design

This nationwide register-based observational cohort study identified all individuals with an AF diagnosis first recorded during hospitalization. Exclusion criteria consisted of unavailable educational attainment; age <30 years, to ensure reliable classification of the highest education level attained; and age > 85 years at AF diagnosis. Morbidity endpoint data were available through December 31, 2009.[12]

We categorized the cohort into three groups based on highest education level attained according to the International Standard Classification of Education (ISCED): primary (ISCED 1–2), secondary (ISCED 3–4), and academic (ISCED 5–8) education. [16,17] Outcomes were a first hospitalization for HF, AMI, or stroke during follow-up. Follow-up started on the date of the index hospitalization and continued for up to five years, with censoring at death, emigration, or the end of the study period, December 31, 2009.

Patients who died within 30 days following the index hospitalization were excluded from the main survival analyses to reduce the influence of deaths that were unlikely to reflect a consequence of AF. A sensitivity analysis was conducted that included patients who died within 30 days of the index hospitalization. To define incident outcomes, endpoint-specific analyses excluded patients with a record of the respective diagnosis prior to the index hospitalization, using all available National Patient Registry data from 1987 to 1994 (the earliest period with nationwide ICD-coded data). Thus, individuals with prior HF, AMI, or stroke were excluded from the respective endpoint analyses. Comorbidity burden was characterized using the Charlson Comorbidity Index (CCI), which summarizes comorbidity-related mortality risk.[18,19] The ICD-9 and ICD-10 codes for CCI conditions (Supplementary Table 2) were defined according to Quan et al.[20] The comorbidity burden was categorized as low (CCI 0–2), moderate (CCI 3–4), or high (CCI ≥ 5), consistent with the previously described dataset.[12] We calculated the CHA_2_DS_2_-VA score to describe thromboembolic risk.[21,22] (Table 1)

**Table 1.**
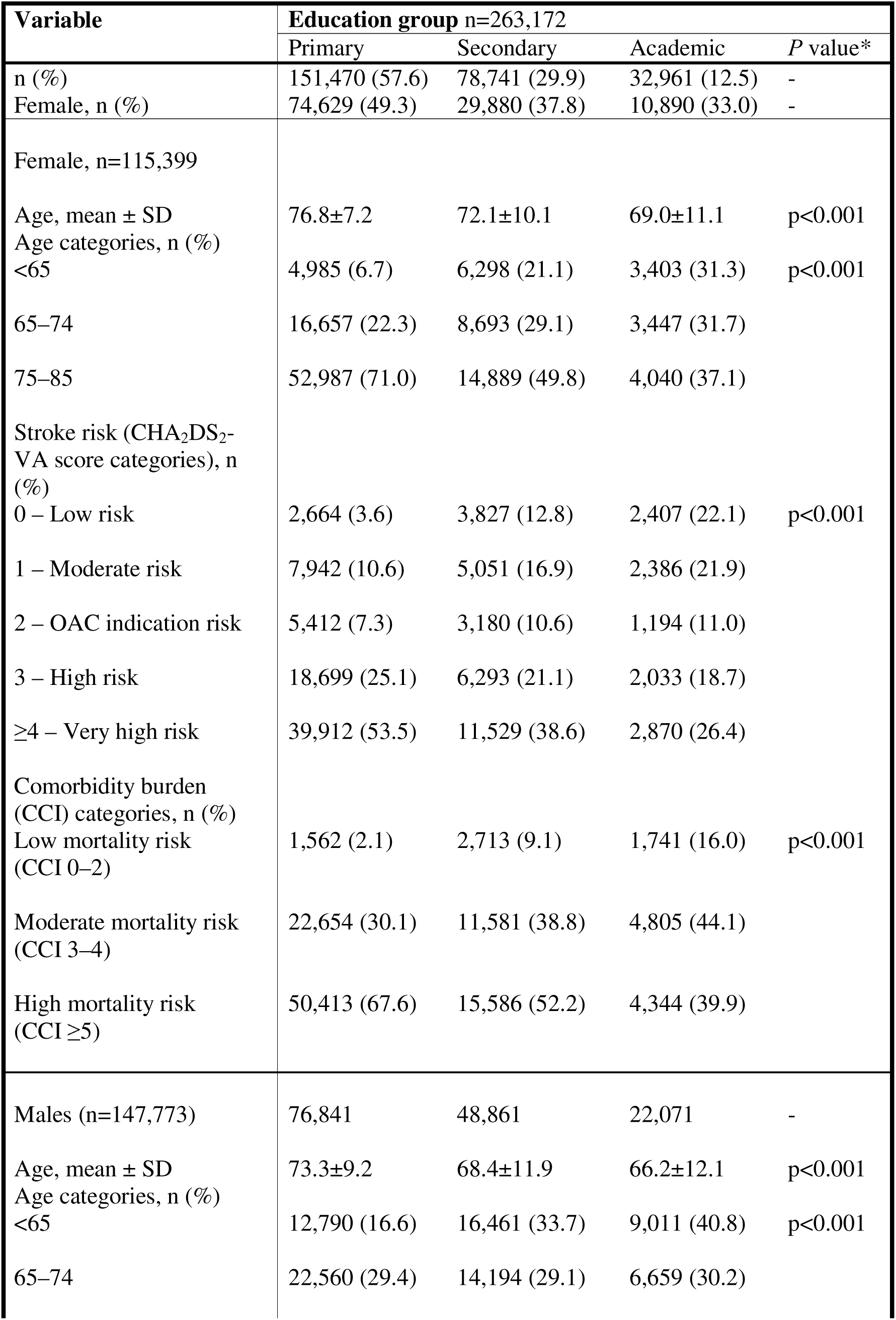
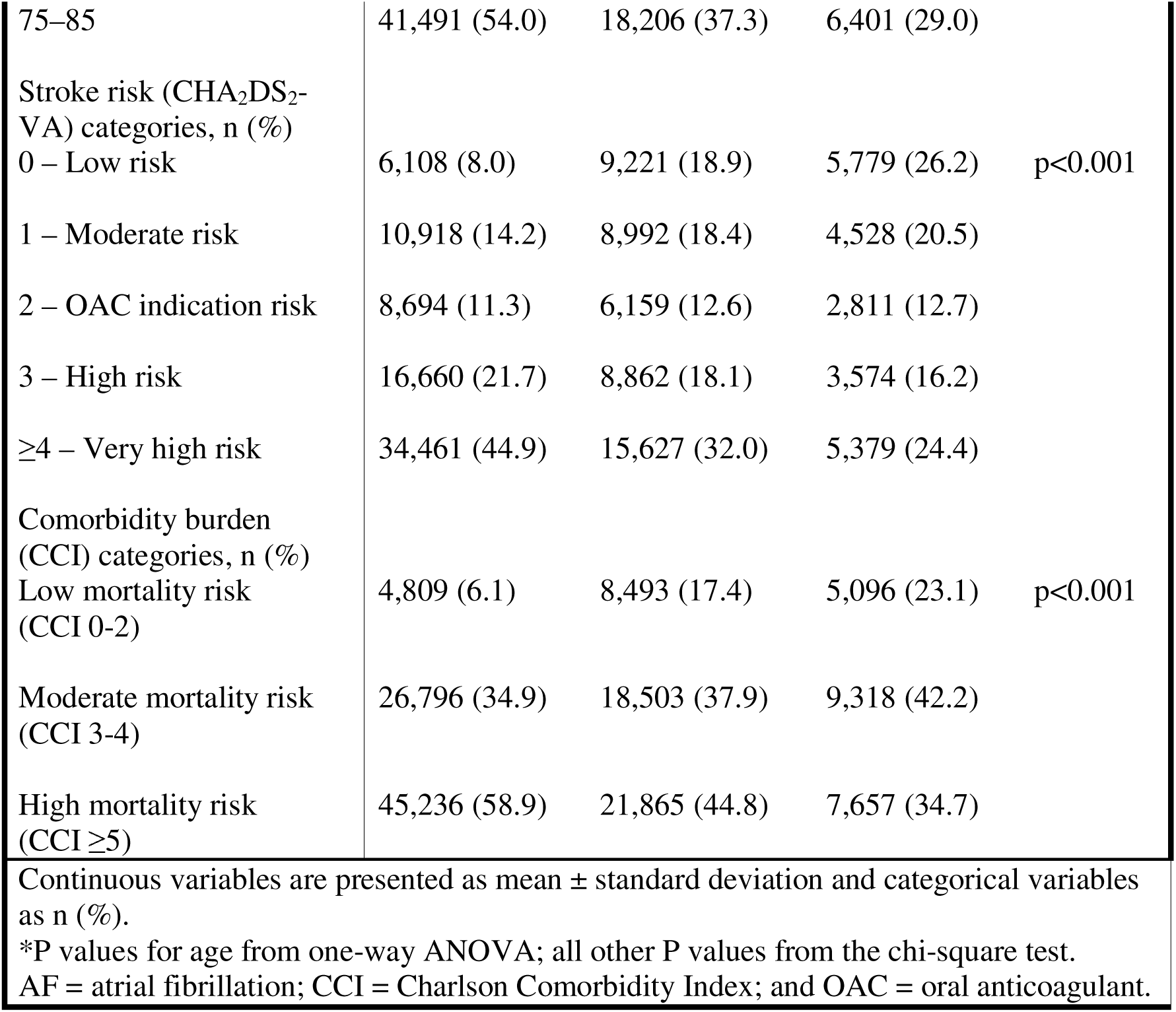
Baseline characteristics of the study population by educational attainment.

### Statistical Analyses

Categorical variables were presented as percentages, and continuous variables as mean ± standard deviation (SD). Differences in the incidence of registered comorbidities across education groups were assessed using the Chi-square test, and age was compared using one-way analysis of variance (ANOVA). Unadjusted cumulative incidence of the endpoint events across education groups was illustrated using Kaplan-Meier failure plots. Cox regression models were used to compare risks across education groups for females and males separately. Results are reported as hazard ratios (HR) with 95% confidence intervals (CI). Analyses were performed unadjusted and adjusted for age, calendar year of AF diagnosis (1995–1999; 2000–2004; 2005–2008), AF as the primary or secondary diagnosis at the index hospitalization, and comorbidity burden using CCI comorbidity diagnosis and hypertension as binary variables. Age was modelled using restricted cubic splines with four knots, according to Harrell.[23] [24] If the proportional hazard assumption was violated for a variable, stratified Cox regression models were used. Non-proportionality was assessed using Schoenfeld residuals (STATA PHTEST). All analyses were performed using STATA versions 16 and 17 (StataCorp, College Station, TX, USA).

### Ethics

The study was conducted in accordance with the Declaration of Helsinki and approved by the Regional Ethical Review Board in Uppsala, Sweden (Dnr: 2009/273). Data were anonymized by the Swedish National Board of Health and Welfare and Statistics Sweden before delivery to the researchers, and the requirement for informed consent was waived.

## Results

Our database included 272,182 patients with a first recorded diagnosis of AF during hospitalization. Of these, 1,448 were excluded because of age <30 years and 7,562 because data on education level were missing. The final study population comprised 263,172 individuals with a mean age of 72.5±10.4 years, 56.2% of whom were male.

The number of males and females was similar only in the primary education group. The proportion of females decreased as education level increased, comprising 37.8% in the secondary and 33.0% in the academic group. Among females, mean age at diagnosis was 76.8±7.2, 72.1±10.1, and 69.0±11.1 years in the primary, secondary, and academic education groups, respectively. Among males, the corresponding ages were 73.3±9.2, 68.4±11.9, and 66.2±12.1 years (Table 1).

Heart failure, AMI, or stroke occurred in 34,243, 25,908, and 21,708 individuals during follow-up, respectively, accounting for 18.1%, 11.8%, and 10.1%, with the majority in the first year following diagnosis. All three outcomes were more common among individuals with lower educational attainment (Figure 1). Females with primary education had higher unadjusted incidence of HF and stroke than males, while incidence of stroke was greater in males in the secondary and academic education groups. For AMI, incidence was higher in males than in females across all education groups.

**Figure 1.a.**
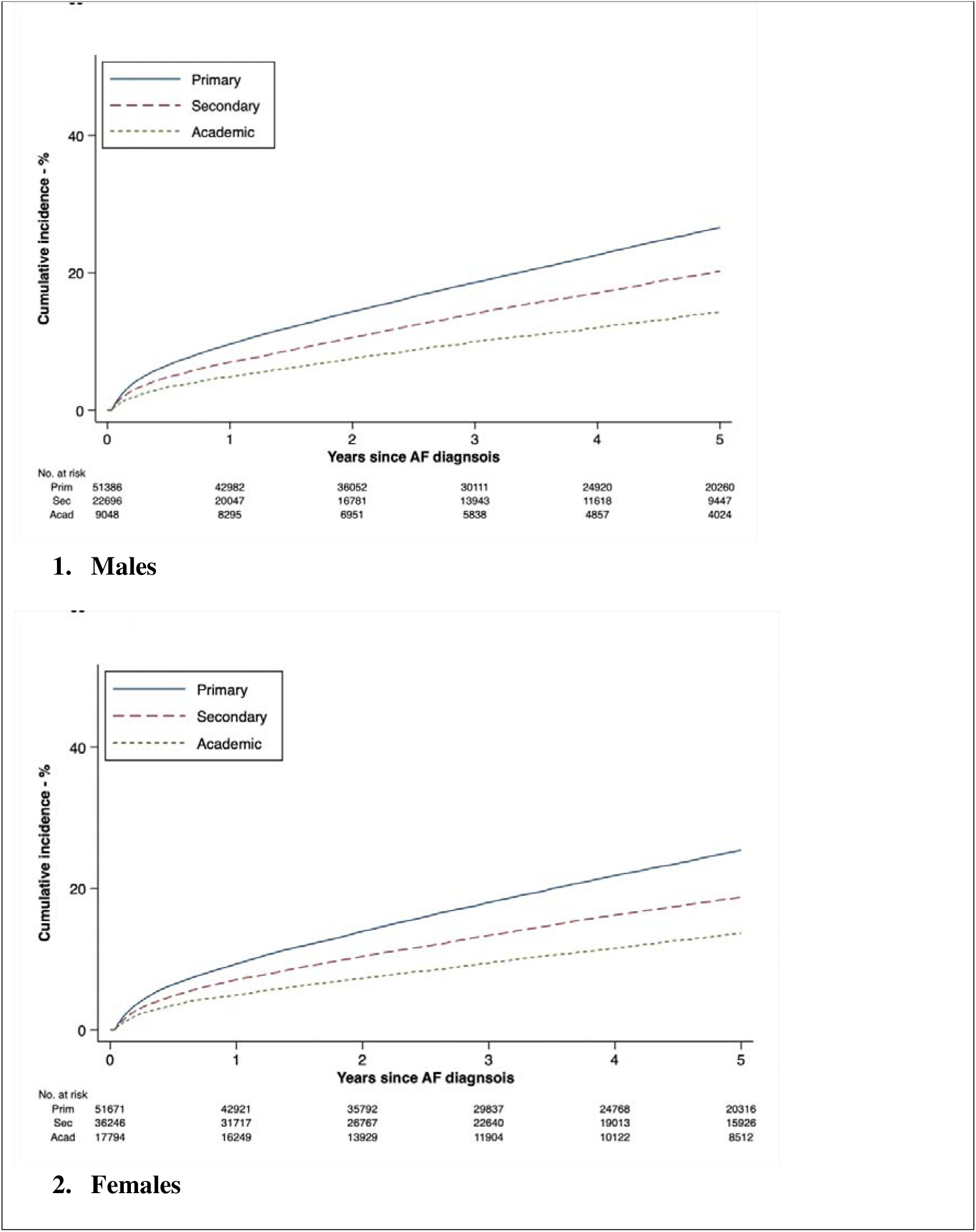
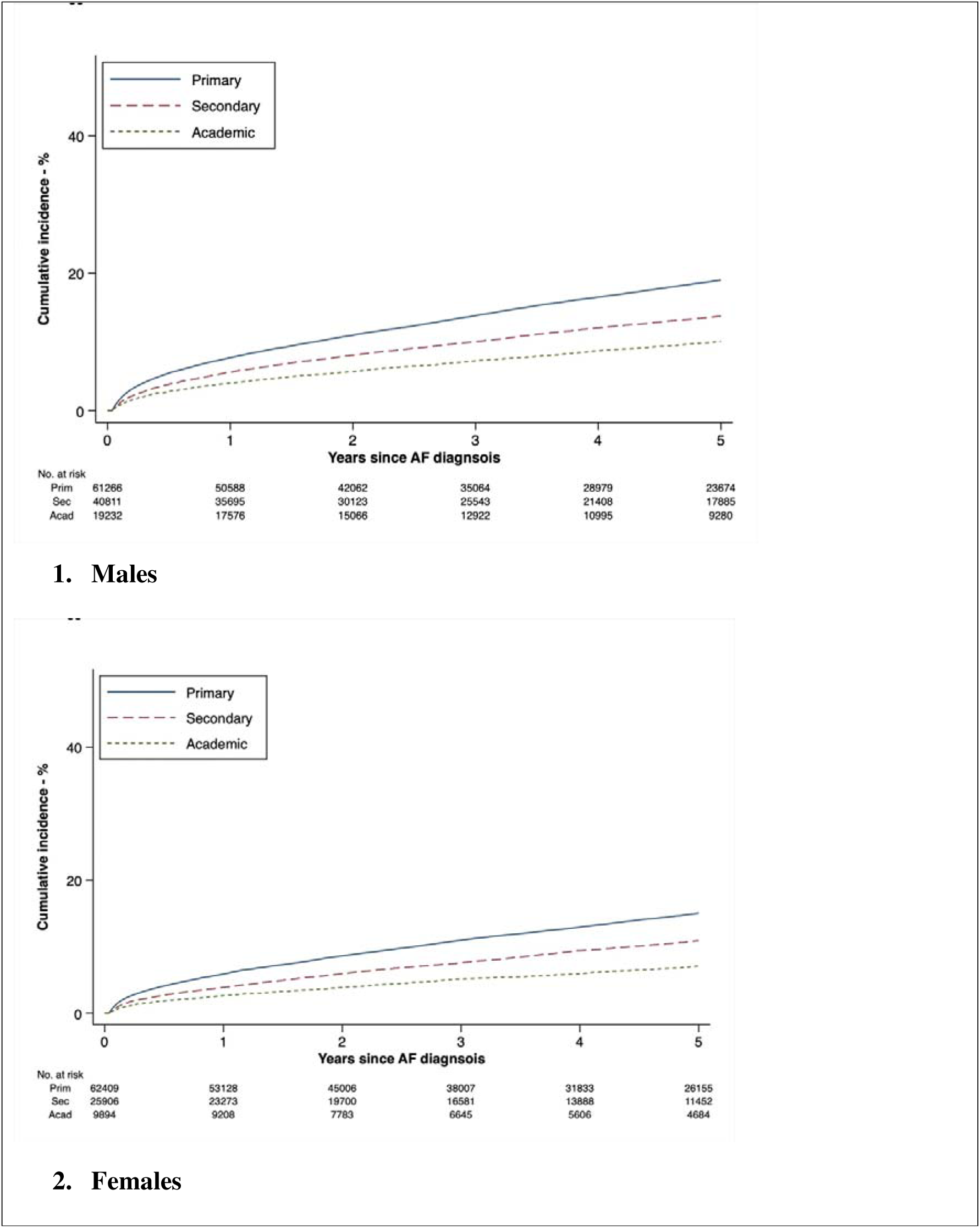
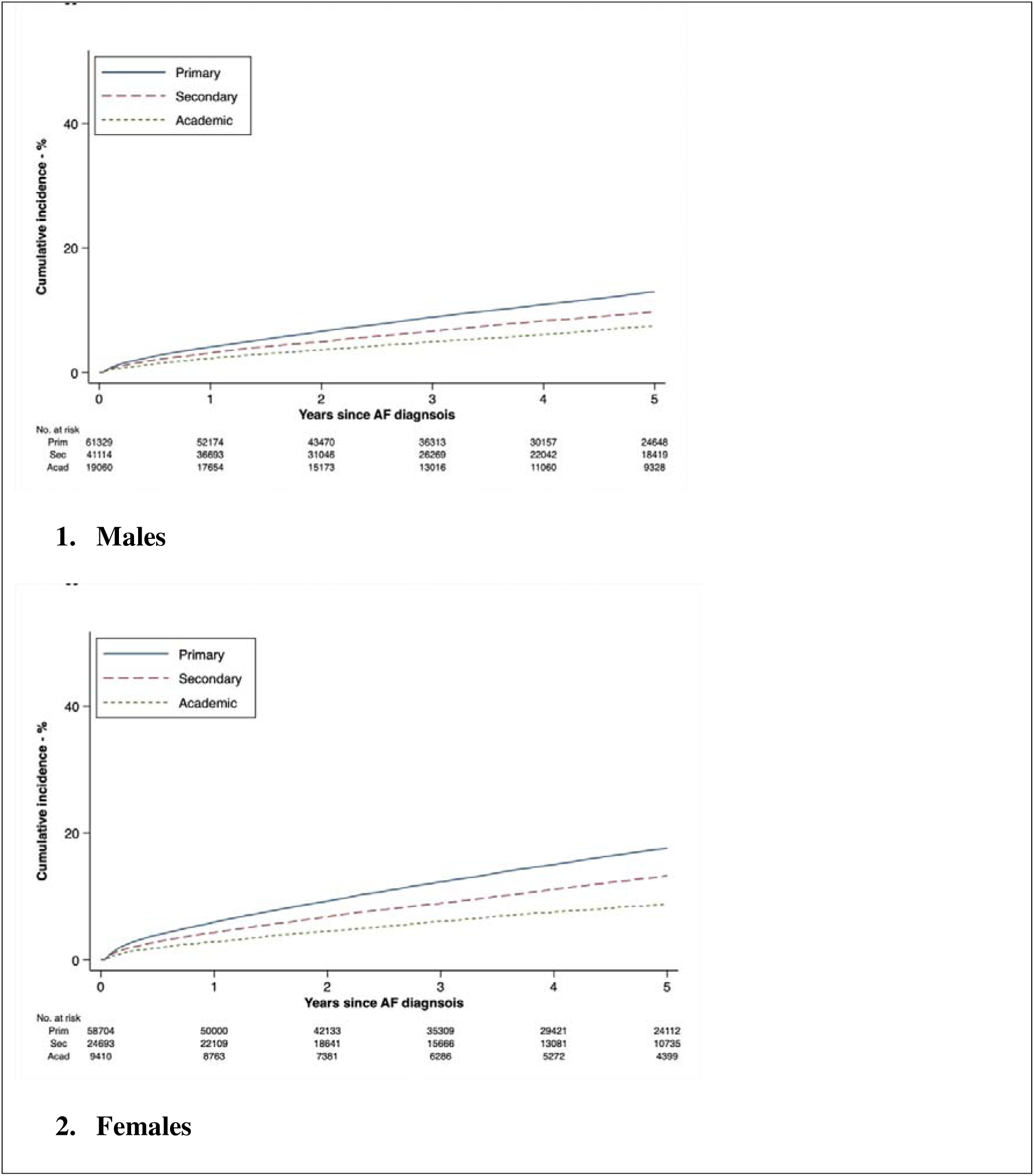
Kaplan-Meier failure estimates for incident heart failure. The y-axis maximum is 0.5 in all panels. **Figure 1.b.** Kaplan-Meier failure estimates for incident acute myocardial infarction Y-axis maximum is 0.5 in all panels. **Figure 1.c.** Kaplan-Meier failure estimates for incident stroke Y-axis maximum is 0.5 in all panels.

Higher educational attainment was associated with a lower risk of HF and AMI (Figure 2). For HF, adjusted HRs for secondary versus primary education were 0.96 (95% CI 0.93–1.00) in females and 0.93 (95% CI 0.90–0.96) in males; for academic versus primary education, HRs were 0.82 (95% CI 0.77–0.87) in females and 0.76 (95% CI 0.72–0.80) in males (Table 2). Sensitivity analyses including patients who died within 30 days following index hospitalization showed no difference in the results (Supplementary Tables 6 and 7).

**Figure 2.a.**
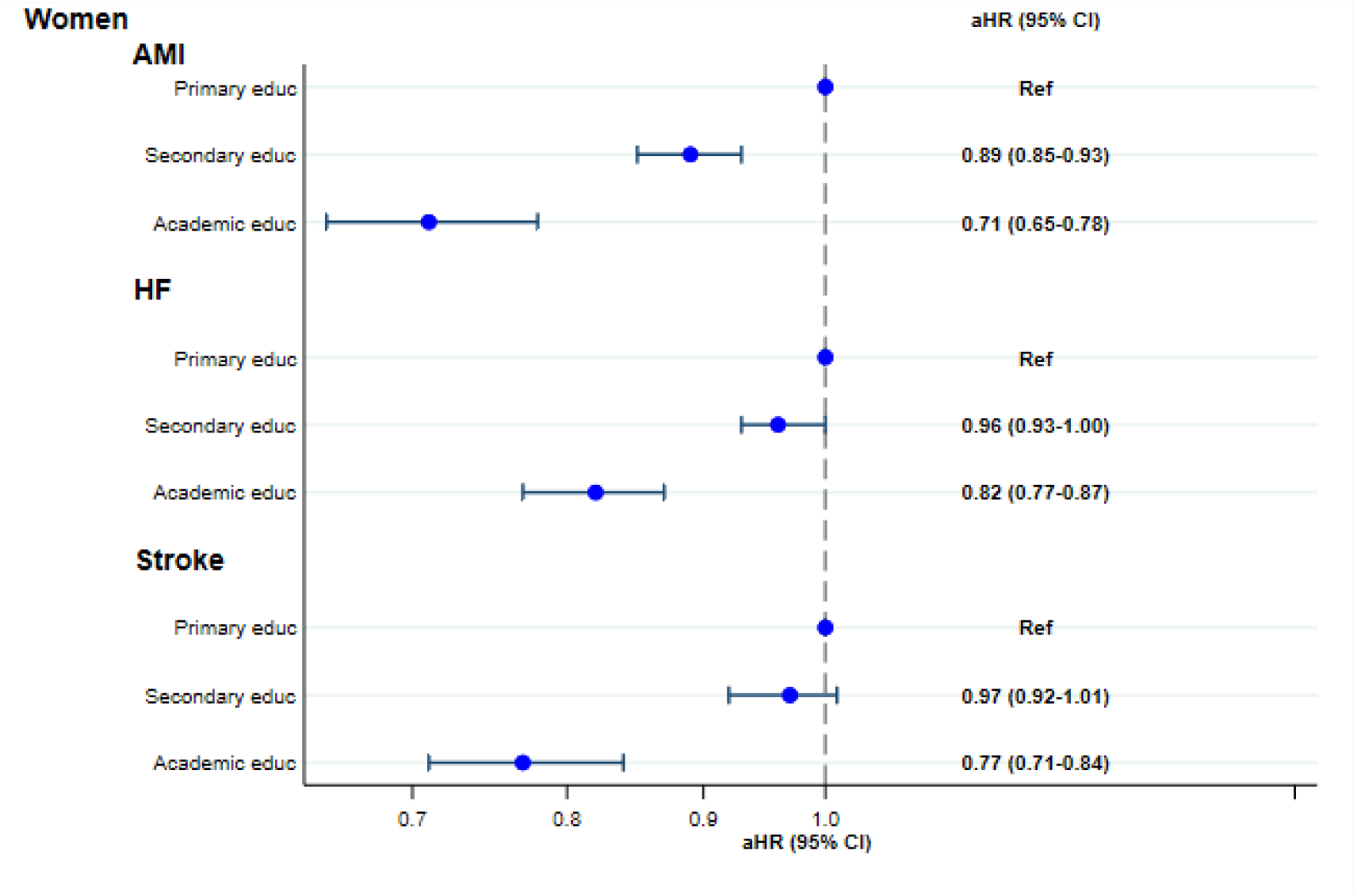
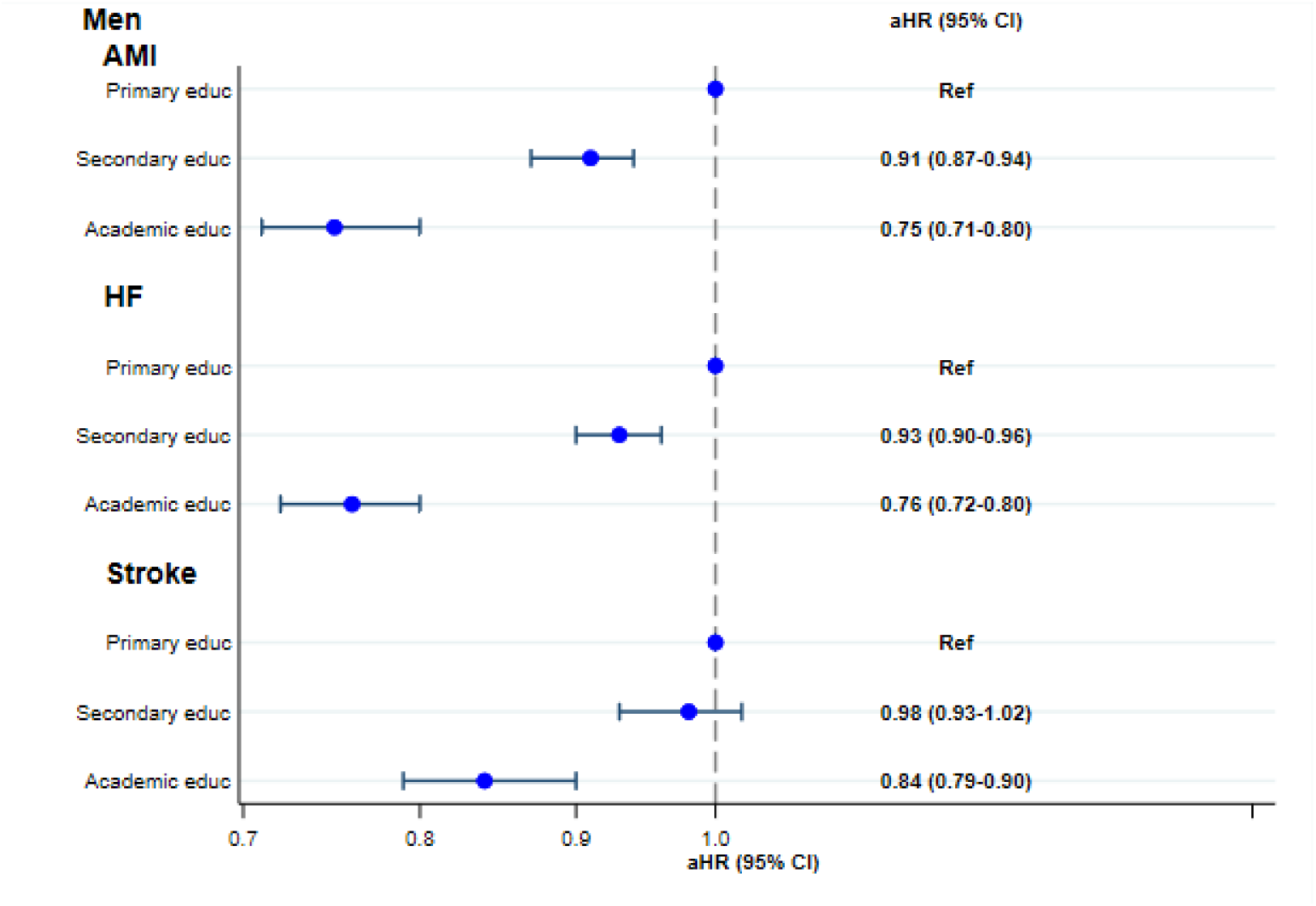
Forest plots of Hazard Ratios for risk of endpoint events by educational attainment, females. **Figure 2.b.** Forest plots of Hazard Ratios for risk of endpoint events by educational attainment, males.

**Table 2.** Five-year risk of heart failure by educational attainment using Cox regression.

| *Females*<br>(*n*=83,130) | *n* | *Events, n (%)* | *Rates \** | *Unadjusted*<br>*HR (95% CI)* | *Adjusted †*<br>*HR (95% CI)* |
| --- | --- | --- | --- | --- | --- |
| *Education group* |  |  |  |  |  |
| Primary | 51,386 | 11,170 (21.7) | 66.2 | Ref | Ref |
| Secondary | 22,696 | 3,748 (16.5) | 48.0 | 0.73 (0.70–0.76) | 0.96 (0.93–1.00) |
| Academic | 9,048 | 1,059 (11.7) | 32.7 | 0.50 (0.47–0.53) | 0.82 (0.77–0.87) |
| *Males*<br>(*n*=105 711) | *n* | *Events, n (%)* | *Rates\** | *Unadjusted*<br>*HR (95% CI)* | *Adjusted †*<br>*HR (95% CI)* |
| *Education group* |  |  |  |  |  |
| Primary | 51,671 | 10,625 (20.6) | 63.2 | Ref | Ref |
| Secondary | 36,246 | 5,614 (15.5) | 44.7 | 0.71 (0.69–0.74) | 0.93 (0.90–0.96) |
| Academic | 17,794 | 2,027 (11.4) | 31.2 | 0.50 (0.48–0.53) | 0.76 (0.72–0.80) |
| *Crude rates are per 1,000 person-years.<br>† Adjusted for age (restricted cubic splines); calendar period of AF diagnosis (1995–1999, 2000–2004, 2005–2008); AF as the primary or secondary admission diagnosis, and baseline comorbidities as defined in the dataset. Stratified Cox regression was used if non-proportional hazards were detected.<br>AF = atrial fibrillation; CI = confidence interval; HR = hazard ratio. |  |  |  |  |  |

For AMI, adjusted HRs for secondary versus primary education were 0.89 (95% CI 0.85–0.93) in females and 0.91 (95% CI 0.87–0.94) in males; for academic versus primary education, adjusted HRs were 0.71 (95% CI 0.65–0.77) in females and 0.75 (95% CI 0.71–0.80) in males (Table 3). Sensitivity analyses showed similar results (Supplementary Tables 6 and 7).

**Table 3.**
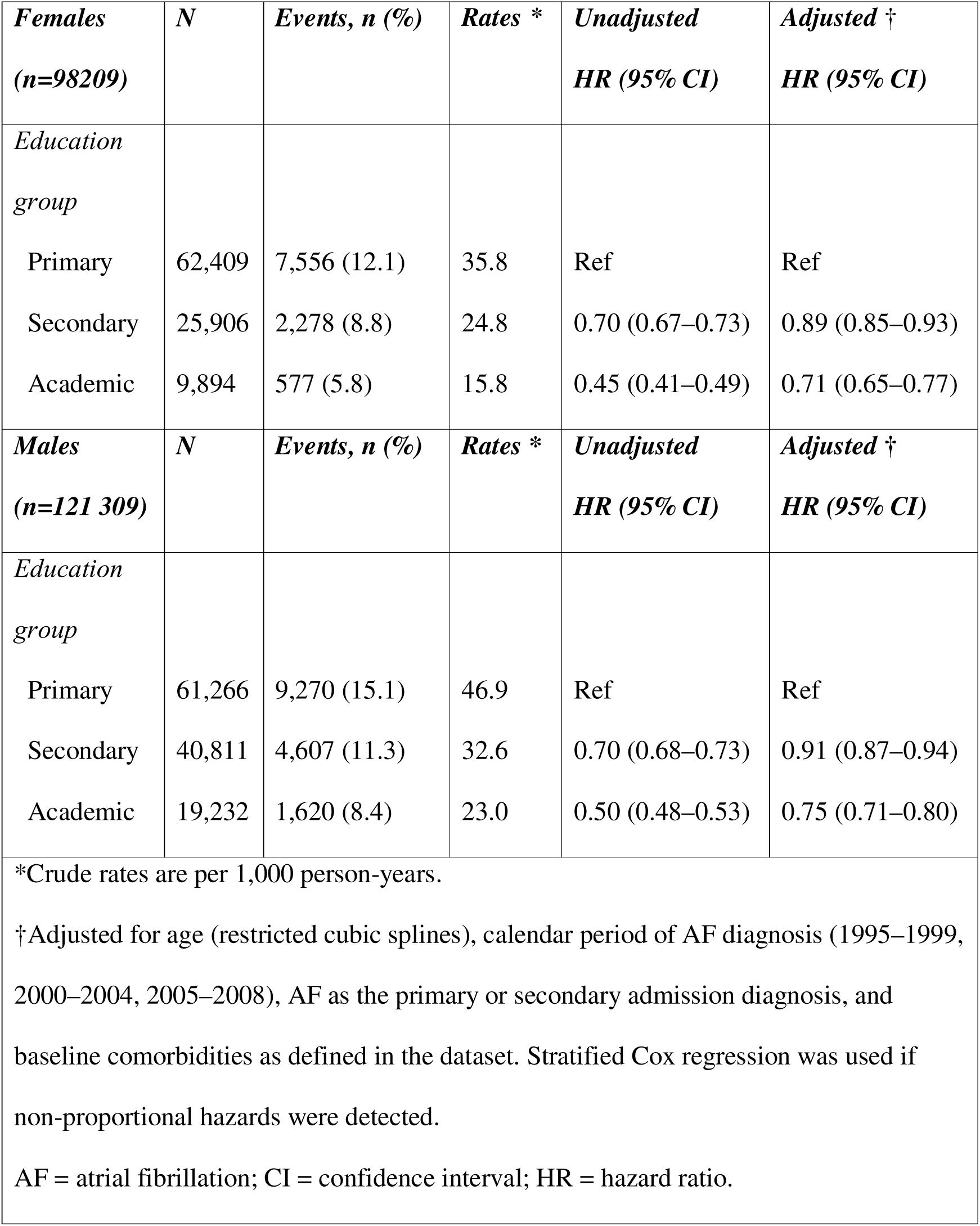
Five-year risk of acute myocardial infarction by educational attainment using Cox regression.

| *Females*<br>*(n=98209)* | *N* | *Events, n (%)* | *Rates \** | *Unadjusted*<br>*HR (95% CI)* | *Adjusted †*<br>*HR (95% CI)* |
| --- | --- | --- | --- | --- | --- |
| *Education*<br>*group* |  |  |  |  |  |
| Primary | 62,409 | 7,556 (12.1) | 35.8 | Ref | Ref |
| Secondary | 25,906 | 2,278 (8.8) | 24.8 | 0.70 (0.67–0.73) | 0.89 (0.85–0.93) |
| Academic | 9,894 | 577 (5.8) | 15.8 | 0.45 (0.41–0.49) | 0.71 (0.65–0.77) |
| *Males*<br>*(n=121 309)* | *N* | *Events, n (%)* | *Rates \** | *Unadjusted*<br>*HR (95% CI)* | *Adjusted †*<br>*HR (95% CI)* |
| *Education*<br>*group* |  |  |  |  |  |
| Primary | 61,266 | 9,270 (15.1) | 46.9 | Ref | Ref |
| Secondary | 40,811 | 4,607 (11.3) | 32.6 | 0.70 (0.68–0.73) | 0.91 (0.87–0.94) |
| Academic | 19,232 | 1,620 (8.4) | 23.0 | 0.50 (0.48–0.53) | 0.75 (0.71–0.80) |
| *Crude rates are per 1,000 person-years.<br>†Adjusted for age (restricted cubic splines), calendar period of AF diagnosis (1995–1999, 2000–2004, 2005–2008), AF as the primary or secondary admission diagnosis, and baseline comorbidities as defined in the dataset. Stratified Cox regression was used if non-proportional hazards were detected.<br>AF = atrial fibrillation; CI = confidence interval; HR = hazard ratio. |  |  |  |  |  |

The association with stroke was significant only in the academic education group: adjusted HR 0.77 (95% CI 0.71–0.83) in females and 0.84 (95% CI 0.79–0.90) in males (Table 4; Figure 2). Results were unchanged in the sensitivity analyses (Supplementary Tables 6 and 7).

**Table 4.** Five-year risk of stroke by educational attainment using Cox regression.

| ***Females***<br>***(n=92,807)*** | ***N*** | ***Events, n (%)*** | ***Rates \**** | ***Unadjusted***<br>***HR (95% CI)*** | ***Adjusted †***<br>***HR (95% CI)*** |
| --- | --- | --- | --- | --- | --- |
| ***Education***<br>***group*** |  |  |  |  |  |
| Primary | 58,704 | 8 165 (13.9) | 41.4 | Ref | Ref |
| Secondary | 24,693 | 2 601 (10.5) | 30.0 | 0.73 (0.70–0.76) | 0.97 (0.93–1.01) |
| Academic | 9,410 | 669 (7.1) | 19.4 | 0.47 (0.44–0.51) | 0.77 (0.71–0.83) |
| ***Males***<br>***(n=121,503)*** | ***n*** | ***Events, n (%)*** | ***Rates \**** | ***Unadjusted***<br>***HR (95% CI)*** | ***Adjusted †***<br>***HR (95% CI)*** |
| ***Education***<br>***group*** |  |  |  |  |  |
| Primary | 61,329 | 6 001 (9.8) | 29.4 | Ref | Ref |
| Secondary | 41,114 | 3 139 (7.6) | 21.6 | 0.74 (0.71–0.77) | 0.98 (0.93–1.02) |
| Academic | 19,060 | 1 133 (5.9) | 16.0 | 0.55 (0.52–0.59) | 0.84 (0.79–0.90) |
| *Crude rates are per 1,000 person-years.<br>†Adjusted for age (restricted cubic splines), calendar period of AF diagnosis (1995–1999, 2000–2004, 2005–2008), AF as the primary or secondary admission diagnosis, and baseline comorbidities as defined in the dataset. Stratified Cox regression was used if non-proportional hazards were detected.<br>AF = atrial fibrillation; CI = confidence interval; HR = hazard ratio. |  |  |  |  |  |

Across education groups, females had higher CHA=DS=-VA scores and CCI scores than males (Table 1). The proportion of females with very high thromboembolic risk (CHA_2_DS_2_-VA ≥ 4) was 53.5%, 38.6%, and 26.4% in the primary, secondary, and academic education groups, respectively, compared with 44.9%, 32.0%, and 24.4% among males. The proportion of patients with high comorbidity burden (CCI ≥ 5) was 67.6%, 52.2%, and 39.9% among females and 58.9%, 44.8%, and 34.7% among males in the primary, secondary, and academic education groups, respectively.

## Discussion

Education beyond the primary level was associated with lower risk of subsequent incident HF and AMI, whereas the association with stroke remained significant only in the academic education group.

The majority of HF, AMI, and stroke events occurred during the first year of follow-up. Although patients were defined by a first recorded AF diagnosis during hospitalization, this may not have represented their first-ever AF episode. Nevertheless, it is reasonable to assume that a substantial proportion had incident or recent-onset AF, and that early initiation of treatment for AF and comorbidities may have contributed to improved risk factor control and delayed progression of AF and/or associated cardiovascular risk.

Heart failure, AMI, and stroke share modifiable cardiovascular risk factors, such as hypertension, smoking, and sedentary lifestyle, which are more prevalent among groups with lower educational attainment.[25] These factors may have contributed to the observed associations of educational attainment and morbidity. Differences in health literacy, access to care, and adherence to treatment recommendations may also have contributed, particularly for outcomes that are sensitive to risk factor control and secondary prevention.[1]

Atrial fibrillation and HF share pathophysiological pathways involving atrial and ventricular remodelling, and the conditions may exert reciprocal effects on disease development and progression.[26] [27] The high incidence of HF observed in our cohort underscores the clinical importance of early identification and management of HF risk factors among patients with AF.[26] Trials of rhythm-control strategies in selected HF populations, including catheter ablation, suggest potential benefits in some patients; however, these findings may not be generalizable to an unselected nationwide AF population and do not explain the education gradient observed in our study.[28], [29]

The association between AF and the risk of incident AMI showed a similar pattern. Atrial fibrillation may be linked to AMI through shared risk factors such as hypertension, diabetes, and dyslipidaemia that promote systemic inflammation, atherosclerosis, and acute ischemic events. Tachyarrhythmia episodes may also cause myocardial ischemia and type 2 myocardial infarction and, rarely, coronary embolism during AF.[30] Conversely, AMI and coronary artery disease are associated with a higher risk of AF, creating a feed-forward loop that may increase morbidity.[31] This elevated risk was also observed in a retrospective observational study from Olmsted County, Minnesota that followed 2,768 patients with newly diagnosed AF for six years.[32] The 5-year unadjusted cumulative incidence of AMI was 14.5% in males and 14.3% in females, compared with 12.8% and 10.6% in our study, respectively. The higher incidence in the Olmsted study may partly reflect differences in endpoint definitions, as incident angina pectoris and unstable angina were also included. In that study, sex differences in incidence became more apparent after adjustment for age.

The association between educational attainment and stroke risk in our study was weakened after adjusting for comorbidities, which may suggest that age and clinical risk factors account for a larger proportion of the observed gradient for stroke than for HF and AMI. An Australian prospective observational cohort study reported a similar inverse relationship of education level and stroke risk in individuals with and without AF.[33] The authors described a non-linear association, with the lowest stroke risk observed in the highest education group. Interaction analyses indicated that lifestyle factors, psychological distress, and concomitant morbidity contributed substantially to the elevated risk. A Mendelian randomization study in populations of European ancestry also suggested an association between genetically-predicted educational attainment and stroke risk, with mediation through cardiovascular risk factors such as dyslipidaemia, hypertension, diabetes, and smoking.[34] These findings support the interpretation that the influence of modifiable risk factors on stroke may differ from their effects on HF and AMI in the AF population.

In addition, socioeconomic differences may affect the initiation and adherence to oral anticoagulation, the cornerstone of stroke prevention in AF, and thereby further contribute to disparities. A Finnish nationwide observational study reported significant differences in initiation of anticoagulation therapy among patients with AF, with adherence approximately 20% higher in the highest versus the lowest education group.[35]

While not formally comparing females and males, we observed that females had higher calculated stroke and comorbidity risk scores, which may be explained by their older age at hospitalization.[36,37] Consistent with these higher risk scores, females experienced higher event rates of HF and stroke. These findings align with a meta-analysis of sex differences in cardiovascular morbidity and mortality, which reported that AF confers a greater relative risk in females than in males.[38]

This large nationwide patient population provides statistical power and supports generalizability, although our study was restricted to hospitalized patients. However, there are limitations to consider: The initial recorded diagnosis of AF does not necessarily mean that it was the patient’s first-ever AF episode. We did not have information on subsequent treatments and adherence during follow-up and therefore could not assess how differences in management may have influenced outcomes.

## Conclusions

Among patients hospitalized with a first recorded diagnosis of AF, lower educational attainment was associated with higher risks of incident HF and AMI, whereas the association with stroke was weaker. Recording education level may help identify patients at increased risk who could benefit from closer monitoring and optimized preventive care.

## Declarations

### Ethical approval

The study was approved by the Regional Ethical Review Board in Uppsala, Sweden (Dnr: 2009/273).

### Data Availability

The data that support the findings of this study are available from the corresponding author upon reasonable request.

## Acknowledgements

Automated assistive writing technologies were used only for grammar checking during the writing process.

## Authors’ contribution

All authors meet the criteria for authorship, and their contributions are as follows: *Study concept and design*: D. Poçi, A. Sztaniszláv, A. Björkenheim, A. Magnuson, N. Edvardsson

*Acquisition of data*: D. Poci, A. Sztaniszláv

*Analysis and interpretation of data*: A. Sztaniszláv, A. Björkenheim, A. Magnuson, N. Edvardsson, D. Poçi.

*Drafting of the manuscript*: A. Sztaniszláv, A. Björkenheim, A. Magnuson, N. Edvardsson, D. Poçi.

*Final approval and critical revision of the manuscript for important intellectual content*: D. Poçi, A. Björkenheim, A. Sztaniszláv, A. Magnuson, N. Edvardsson, *Statistical expertise*: A. Magnuson, A. Sztaniszláv

## Supplemental Material

Supplementary Tables 1-7

---

**Supplementary Table 1.** Definitions of atrial fibrillation and outcome events according to ICD-9 and ICD-10.

| Diagnosis | ICD-9 | ICD-10 |
| --- | --- | --- |
| Atrial fibrillation | 427 D (DA, DB, DC, DD, DW) | I48 and I48.9 (A, B, C, D, E, F, P, X) |
| Myocardial infarction | 410 (A, B, W, X), 411 (A, B, C, X), 412 | I21 (0, 1, 2, 3, 4, 9), I22 (0, 1, 8, 9) I25.2 |
| Heart failure | 428 (A, B, X) | I50 (0, 1, 9) |
| Stroke | 433 (A, B, C, D, W, X), 434 (A, B, X), 435 | I63 (0, 1, 2, 3, 4, 5, 6, 8, 9), I64 and G45 (0, 1, 2, 3, 4, 8, 9) |

**Supplementary Table 2.** Charlson comorbidity index comorbidities included in the statistical analysis.

|  |  |
| --- | --- |
| Diabetes mellitus | Chronic obstructive pulmonary disease |
| Acute myocardial infarction | Peripheral artery disease |
| Coronary artery disease | Dementia |
| Heart failure | Rheumatic disease |
| Cerebrovascular event | Mild liver disease |
| Cancer | Moderate to severe liver disease |
| Chronic kidney disease | Peptic ulcer |

**Supplementary Table 3.**
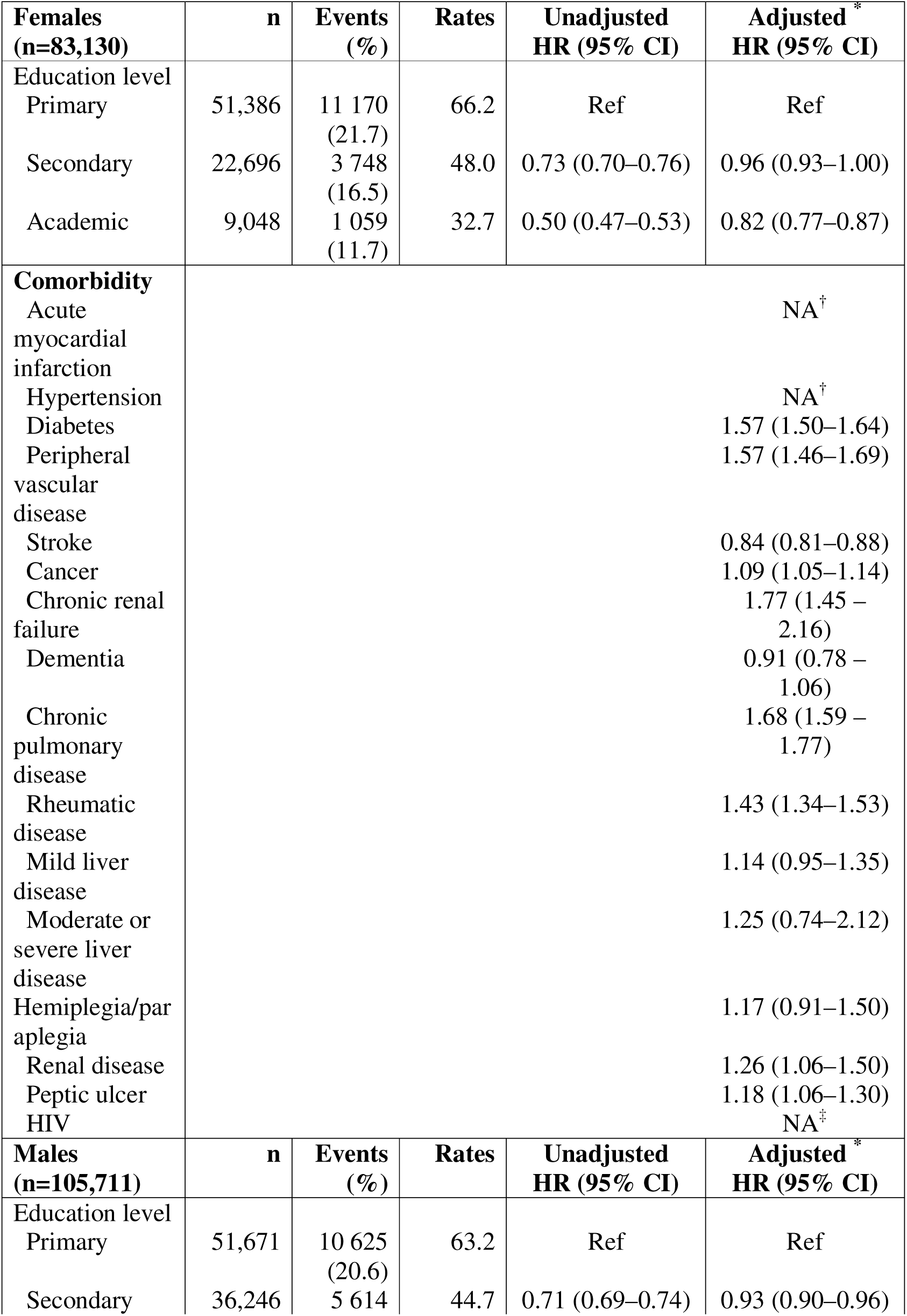
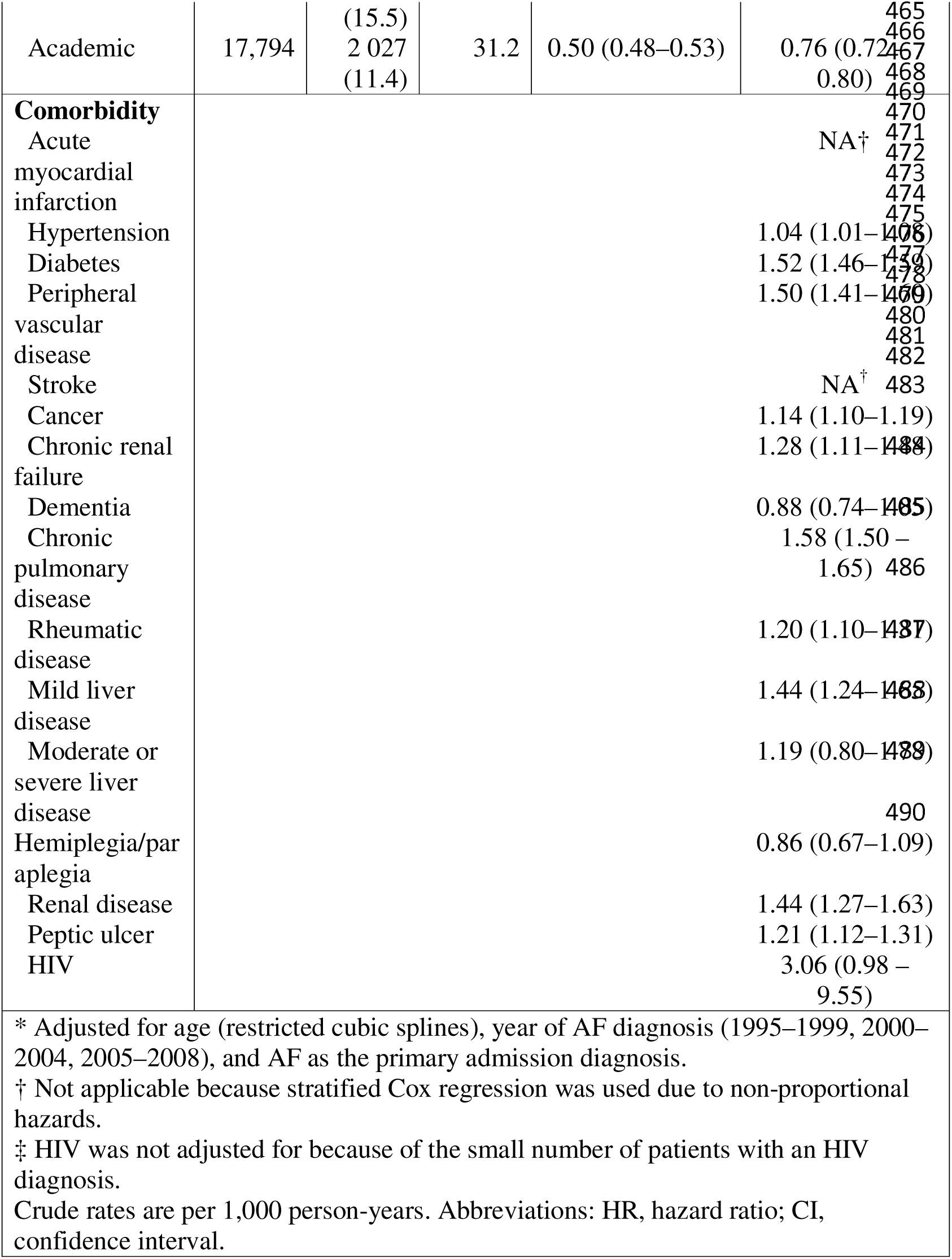
Comparison of five-year heart failure risk across education levels using Cox regression.

**Supplementary Table 4.**
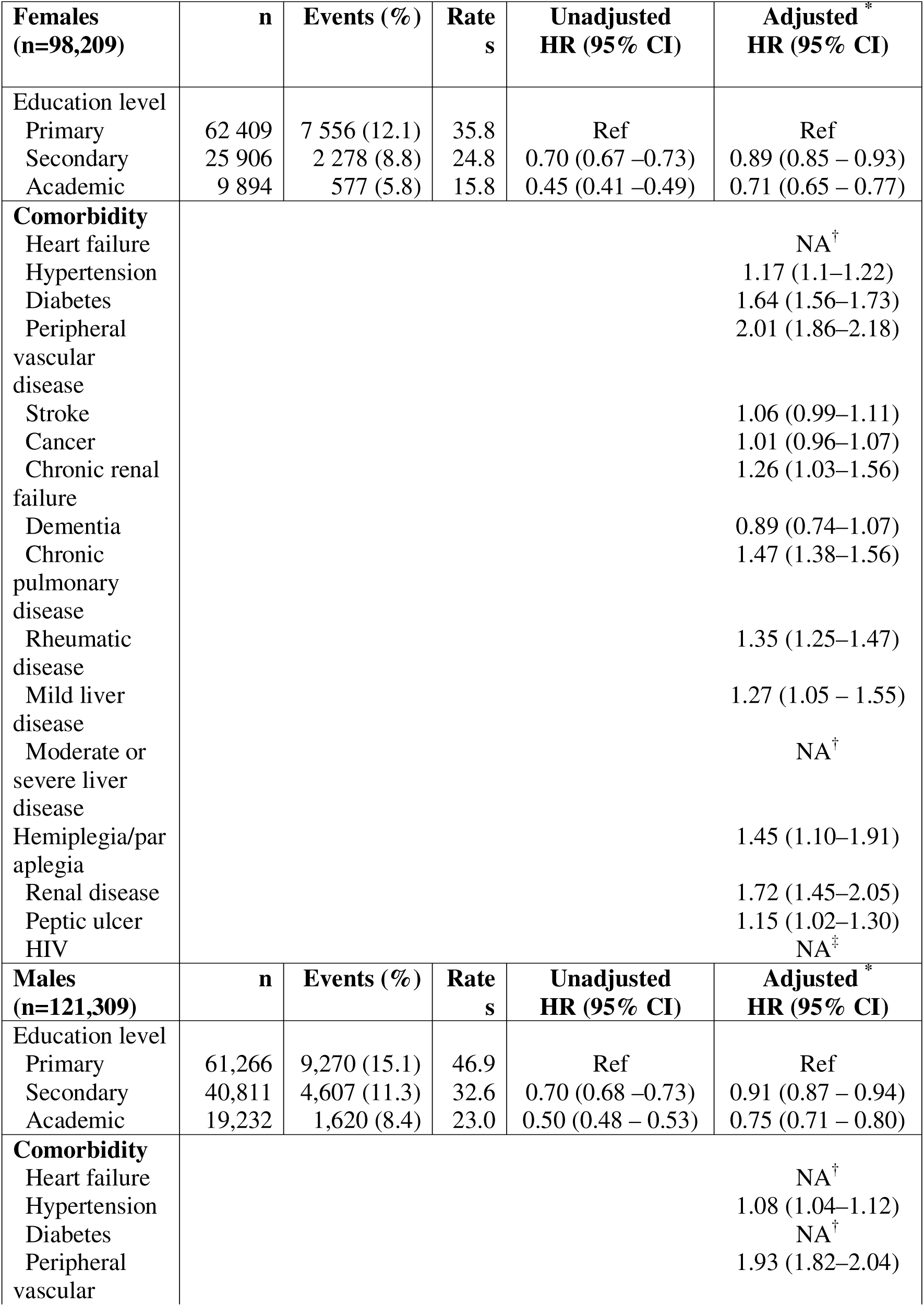
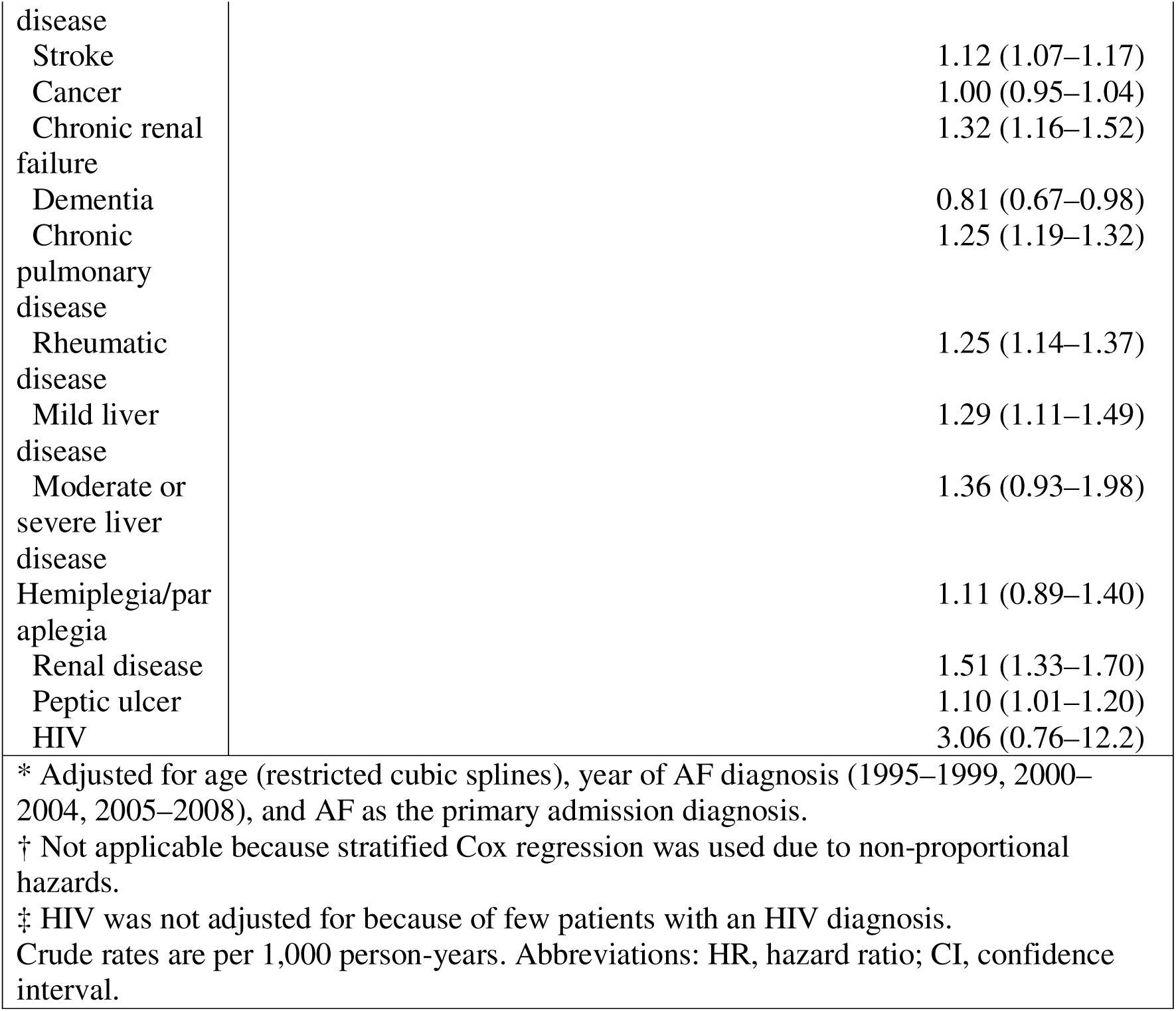
Comparison of five-year risk of acute myocardial infarction across education levels based on Cox regression.

| **Females**<br>**(n=98,209)** | **n** | **Events (%)** | **Rates** | **Unadjusted**<br>**HR (95% CI)** | **Adjusted\***<br>**HR (95% CI)** |
| --- | --- | --- | --- | --- | --- |
| Education level |  |  |  |  |  |
| Primary | 62 409 | 7 556 (12.1) | 35.8 | Ref | Ref |
| Secondary | 25 906 | 2 278 (8.8) | 24.8 | 0.70 (0.67 –0.73) | 0.89 (0.85 – 0.93) |
| Academic | 9 894 | 577 (5.8) | 15.8 | 0.45 (0.41 –0.49) | 0.71 (0.65 – 0.77) |
| **Comorbidity** |  |  |  |  |  |
| Heart failure |  |  |  |  | NA<sup>†</sup> |
| Hypertension |  |  |  |  | 1.17 (1.1–1.22) |
| Diabetes |  |  |  |  | 1.64 (1.56–1.73) |
| Peripheral<br>vascular<br>disease |  |  |  |  | 2.01 (1.86–2.18) |
| Stroke |  |  |  |  | 1.06 (0.99–1.11) |
| Cancer |  |  |  |  | 1.01 (0.96–1.07) |
| Chronic renal<br>failure |  |  |  |  | 1.26 (1.03–1.56) |
| Dementia |  |  |  |  | 0.89 (0.74–1.07) |
| Chronic<br>pulmonary<br>disease |  |  |  |  | 1.47 (1.38–1.56) |
| Rheumatic<br>disease |  |  |  |  | 1.35 (1.25–1.47) |
| Mild liver<br>disease |  |  |  |  | 1.27 (1.05 – 1.55) |
| Moderate or<br>severe liver<br>disease |  |  |  |  | NA<sup>†</sup> |
| Hemiplegia/par<br>aplegia |  |  |  |  | 1.45 (1.10–1.91) |
| Renal disease |  |  |  |  | 1.72 (1.45–2.05) |
| Peptic ulcer |  |  |  |  | 1.15 (1.02–1.30) |
| HIV |  |  |  |  | NA<sup>‡</sup> |
| **Males**<br>**(n=121,309)** | **n** | **Events (%)** | **Rates** | **Unadjusted**<br>**HR (95% CI)** | **Adjusted\***<br>**HR (95% CI)** |
| Education level |  |  |  |  |  |
| Primary | 61,266 | 9,270 (15.1) | 46.9 | Ref | Ref |
| Secondary | 40,811 | 4,607 (11.3) | 32.6 | 0.70 (0.68 –0.73) | 0.91 (0.87 – 0.94) |
| Academic | 19,232 | 1,620 (8.4) | 23.0 | 0.50 (0.48 – 0.53) | 0.75 (0.71 – 0.80) |
| **Comorbidity** |  |  |  |  |  |
| Heart failure |  |  |  |  | NA<sup>†</sup> |
| Hypertension |  |  |  |  | 1.08 (1.04–1.12) |
| Diabetes |  |  |  |  | NA<sup>†</sup> |
| Peripheral<br>vascular |  |  |  |  | 1.93 (1.82–2.04) |

|  |  |
| --- | --- |
| disease |  |
| Stroke | 1.12 (1.07–1.17) |
| Cancer | 1.00 (0.95–1.04) |
| Chronic renal failure | 1.32 (1.16–1.52) |
| Dementia | 0.81 (0.67–0.98) |
| Chronic pulmonary disease | 1.25 (1.19–1.32) |
| Rheumatic disease | 1.25 (1.14–1.37) |
| Mild liver disease | 1.29 (1.11–1.49) |
| Moderate or severe liver disease | 1.36 (0.93–1.98) |
| Hemiplegia/paraplegia | 1.11 (0.89–1.40) |
| Renal disease | 1.51 (1.33–1.70) |
| Peptic ulcer | 1.10 (1.01–1.20) |
| HIV | 3.06 (0.76–12.2) |

\* Adjusted for age (restricted cubic splines), year of AF diagnosis (1995–1999, 2000–2004, 2005–2008), and AF as the primary admission diagnosis.
† Not applicable because stratified Cox regression was used due to non-proportional hazards.
‡ HIV was not adjusted for because of few patients with an HIV diagnosis.
Crude rates are per 1,000 person-years. Abbreviations: HR, hazard ratio; CI, confidence interval.

**Supplementary Table 5.**
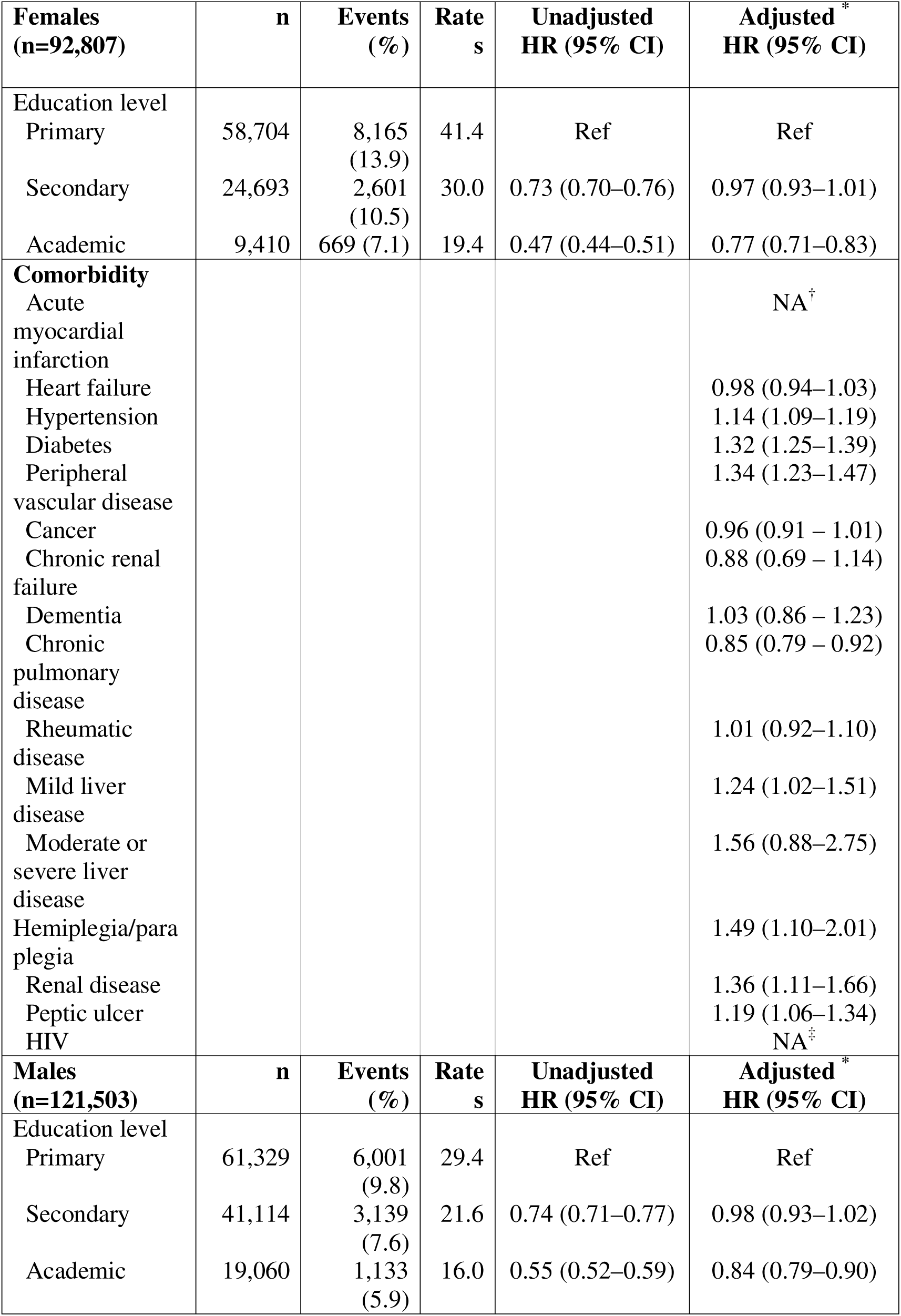
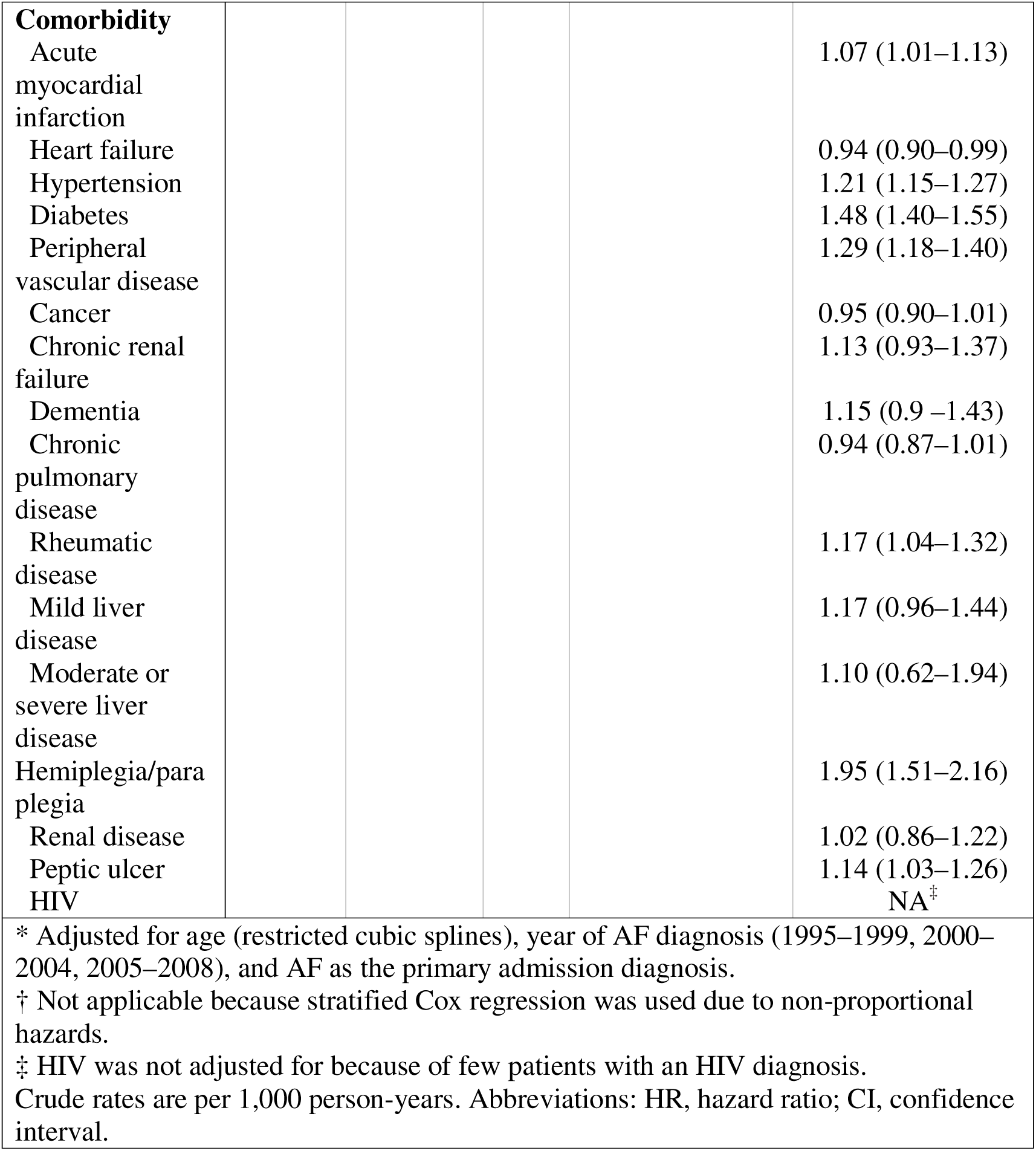
Comparison of Five-year stroke risk across education levels based on Cox regression.

**Supplementary Table 6:**
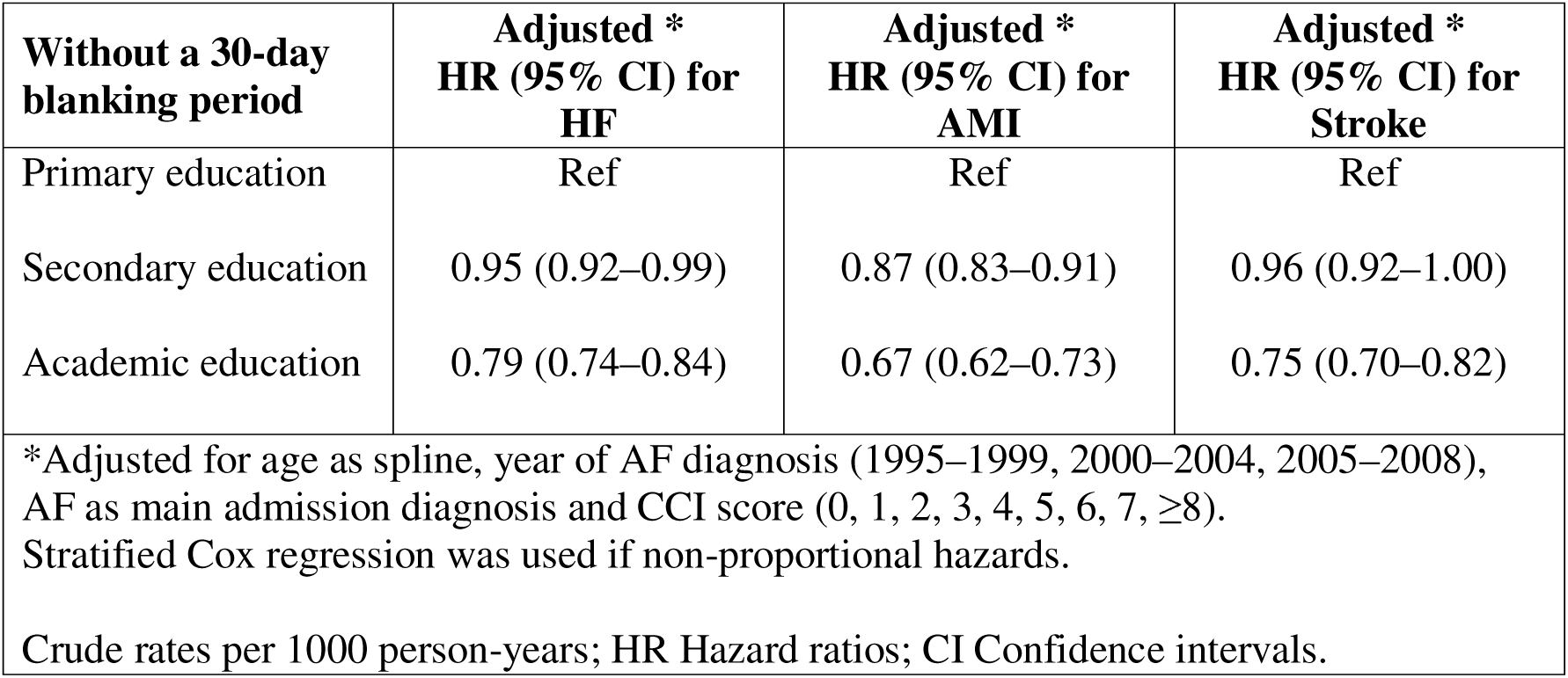
Sensitivity analysis of five-year outcome risk by educational attainment using Cox regression, with and without a 30-day blanking period in females.

| **Without a 30-day blanking period** | **Adjusted \***<br>**HR (95% CI) for**<br>**HF** | **Adjusted \***<br>**HR (95% CI) for**<br>**AMI** | **Adjusted \***<br>**HR (95% CI) for**<br>**Stroke** |
| --- | --- | --- | --- |
| Primary education | Ref | Ref | Ref |
| Secondary education | 0.95 (0.92–0.99) | 0.87 (0.83–0.91) | 0.96 (0.92–1.00) |
| Academic education | 0.79 (0.74–0.84) | 0.67 (0.62–0.73) | 0.75 (0.70–0.82) |
| *Adjusted for age as spline, year of AF diagnosis (1995–1999, 2000–2004, 2005–2008),<br>AF as main admission diagnosis and CCI score (0, 1, 2, 3, 4, 5, 6, 7, ≥8).<br>Stratified Cox regression was used if non-proportional hazards.<br>Crude rates per 1000 person-years; HR Hazard ratios; CI Confidence intervals. |  |  |  |

**Supplementary Table 7:**
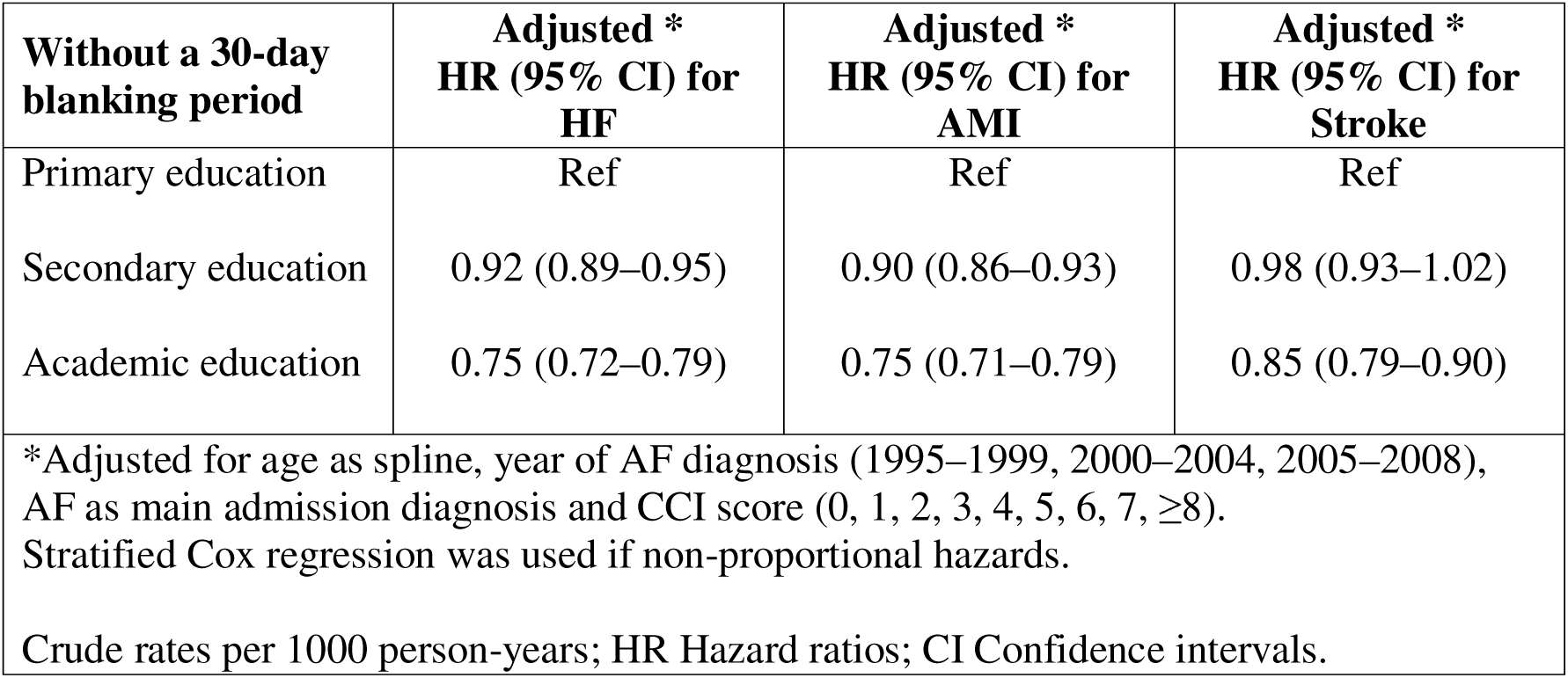
Sensitivity analysis of five-year outcome risk relative to educational attainment based on Cox regression, with and without a 30-day blanking period in males.

## REFERENCES

1. Rangarajan S, Md J, Lamelas P, Teo K, Attaei MA, Boström B, et al. Socioeconomic status and risk of cardiovascular disease in 20 low-income, middle-income, and high-income countries: the Prospective Urban Rural Epidemiologic (PURE) study. Lancet Glob Health. 2019;7:e748–60. 10.1016/S2214-109X(19)30045-2

2. Goli NM, Thompson T, Sears SF, Mounsey JP, Chung E, Schwartz J, et al. Educational attainment is associated with atrial fibrillation symptom severity. PACE - Pacing Clin Electrophysiol. 2012;35:1090–6. 10.1111/j.1540-8159.2012.03482.x

3. Galobardes B, Shaw M, Lawlor DA, Lynch JW, Smith GD. Indicators of socioeconomic position (part 1) [Internet]. J. Epidemiol. Community Health. 2006. p. 7–12. 10.1136/jech.2004.023531

4. Lippi G, Sanchis-Gomar F, Cervellin G. Global epidemiology of atrial fibrillation: An increasing epidemic and public health challenge. Int J Stroke. 2020;0:1–5. 10.1177/1747493019897870

5. Buja A, Rebba V, Montecchio L, Renzo G, Baldo V, Cocchio S, et al. The Cost of Atrial Fibrillation: A Systematic Review. Value Health. 2024;27:527–41. 10.1016/j.jval.2023.12.015

6. Sandhu RK, Wilton SB, Islam S, Atzema CL, Deyell M, Wyse DG, et al. Temporal Trends in Population Rates of Incident Atrial Fibrillation and Atrial Flutter Hospitalizations, Stroke Risk, and Mortality Show Decline in Hospitalizations. Can J Cardiol. 2021;37:310–8. 10.1016/j.cjca.2020.04.026

7. Friberg J, Buch P, Scharling H, Gadsbøll N, Jensen GB. Rising Rates of Hospital Admissions for Atrial Fibrillation: Epidemiology. 2003;14:666–72. 10.1097/01.ede.0000091649.26364.c0

8. Sztaniszláv Á, Björkenheim A, Magnuson A, Bryngelsson I-L, Edvardsson N, Poci D. The impact of education level on all-cause mortality in patients with atrial fibrillation. Sci Rep. 2024;14:25386. 10.1038/s41598-024-74478-2

9. Ngo LTH, Peng Y, Denman R, Yang I, Ranasinghe I. Long-term outcomes after hospitalization for atrial fibrillation or flutter. Eur Heart J. 2024;ehae204. 10.1093/eurheartj/ehae204

10. Kreatsoulas C, Anand SS. The impact of social determinants on cardiovascular disease. Can J Cardiol. 2010;26:8C–13C. 10.1016/S0828-282X(10)71075-8

11. Liao L, Zhuang X, Zhang S, Liao X, Li W. Education and heart failure: New insights from the atherosclerosis risk in communities study and mendelian randomization study. Int J Cardiol. 2021;324:115–21. 10.1016/j.ijcard.2020.09.068

12. Sztaniszlav A, Bjorkenheim A, Magnuson A, Bryngelsson IL, Edvardsson N, Poci D. The role of education level in the mortality of hospitalized patients with atrial fibrillation. Europace. 2023;25:euad122.026. 10.1093/europace/euad122.026

13. Andersson T, Magnuson A, Bryngelsson IL, Frøbert O, Henriksson KM, Edvardsson N, et al. All-cause mortality in 272 186 patients hospitalized with incident atrial fibrillation 1995-2008: A Swedish nationwide long-term case-control study. Eur Heart J. 2013;34:1061–7. 10.1093/eurheartj/ehs469

14. Baturova MA, Lindgren A, Carlson J, Shubik YV, Bertil Olsson S, Platonov PG. Atrial fibrillation in patients with ischaemic stroke in the Swedish national patient registers: how much do we miss? EP Eur. 2014;16:1714–9. 10.1093/europace/euu165

15. Smith JG, Platonov PG, Hedblad B, Engström G, Melander O. Atrial fibrillation in the Malmö diet and cancer study: A study of occurrence, risk factors and diagnostic validity. Eur J Epidemiol. 2010;25:95–102. 10.1007/s10654-009-9404-1

16. UNESCO Institute for Statistics. International Standard Classification of Education (ISCED) 2011 [Internet]. UNESCO Institute for Statistics; 2012 [cited 2026 Mar 23]. 10.15220/978-92-9189-123-8-en

17. Odelholm H. Swedish Standard Classification of Education [Internet]. Örebro, Sweden: Statistics of Sweden; 1996. https://www.scb.se/contentassets/aeeedec0e28c465aa524429407dcd5ba/mis-1996-1.pdf

18. Rashid M, Kwok CS, Gale CP, Doherty P, Olier I, Sperrin M, Kontopantelis E, Peat G MMA, Rashid M, Kwok CS, Gale CP, Doherty P, Olier I, et al. Impact of co-morbid burden on mortality in patients with coronary heart disease, heart failure, and cerebrovascular accident: a systematic review and meta-analysis. Eur Heart J Qual Care Clin Outcomes. England; 2017;3:20–36. 10.1093/ehjqcco/qcw025

19. Mary C, Ronald M. A new method of classifying prognostic comorbidity in longitudinal studies: Developement and validation. J Chronic Dis. 1987;40:373–83. 10.1016/0021-9681(87)90171-8

20. Quan H, Sundararajan V, Halfon P, Fong A, Burnand B, Luthi JC, et al. Coding algorithms for defining comorbidities in ICD-9-CM and ICD-10 administrative data. Med Care. 2005;43:1130–9. 10.1097/01.mlr.0000182534.19832.83

21. Van Gelder IC, Rienstra M, Bunting KV, Casado-Arroyo R, Caso V, Crijns HJGM, et al. 2024 ESC Guidelines for the management of atrial fibrillation developed in collaboration with the European Association for Cardio-Thoracic Surgery (EACTS). Eur Heart J. 2024;ehae176. 10.1093/eurheartj/ehae176

22. Lip GYH, Nieuwlaat R, Pisters R, Lane DA, Crijns HJGM, Andresen D, et al. Refining clinical risk stratification for predicting stroke and thromboembolism in atrial fibrillation using a novel risk factor-based approach: The Euro Heart Survey on atrial fibrillation. Chest. 2009/09/19 ed. 2010;137:263–72. 10.1378/chest.09-1584

23. Helmreich JE. Regression Modeling Strategies with Applications to Linear Models, Logistic and Ordinal Regression and Survival Analysis (2nd Edition). J Stat Softw. 2016;70:3–5. 10.18637/jss.v070.b02

24. Belias M, Rovers MM, Hoogland J, Reitsma JB, Debray TPA, IntHout J. Predicting personalised absolute treatment effects in individual participant data meta-analysis: An introduction to splines. Res Synth Methods. 2022;13:255–83. 10.1002/jrsm.1546

25. Shea S, Stein AD, Basch CE, Freudenheim J, Lantigua R, Maylahn C, et al. Independent Associations of Educational Attainment and Ethnicity with Behavioral Risk Factors for Cardiovascular Disease. Am J Epidemiol. 1991;134:567–82. 10.1093/oxfordjournals.aje.a116130

26. Prabhu S, Voskoboinik A, Kaye DM, Kistler PM. Atrial Fibrillation and Heart Failure — Cause or Effect? Heart Lung Circ. 2017;26:967–74. 10.1016/j.hlc.2017.05.117

27. Börschel CS, Schnabel RB. The imminent epidemic of atrial fibrillation and its concomitant diseases – Myocardial infarction and heart failure - A cause for concern. Int J Cardiol. 2019;287:162–73. 10.1016/j.ijcard.2018.11.123

28. Marrouche NF, Brachmann J, Andresen D, Siebels J, Boersma L, Jordaens L, et al. Catheter Ablation for Atrial Fibrillation with Heart Failure. N Engl J Med. 2018;378:417–27. 10.1056/NEJMoa1707855

29. Simader FA, Howard JP, Ahmad Y, Saleh K, Naraen A, Samways JW, et al. Catheter ablation improves cardiovascular outcomes in patients with atrial fibrillation and heart failure: a meta-analysis of randomized controlled trials. EP Eur. 2023;25:341–50. 10.1093/europace/euac173

30. Violi F, Soliman EZ, Pignatelli P, Pastori D. Atrial Fibrillation and Myocardial Infarction: A Systematic Review and Appraisal of Pathophysiologic Mechanisms. J Am Heart Assoc. 2016;5:e003347. 10.1161/JAHA.116.003347

31. Staerk L, Sherer JA, Ko D, Benjamin EJ, Helm RH. Atrial Fibrillation: Epidemiology, Pathophysiology, and Clinical Outcomes. Circ Res. 2017;120:1501–17. 10.1161/CIRCRESAHA.117.309732

32. Miyasaka Y, Barnes ME, Gersh BJ, Cha SS, Bailey KR, Seward JB, et al. Coronary Ischemic Events after First Atrial Fibrillation: Risk and Survival. Am J Med. 2007;120:357–363.e1. 10.1016/j.amjmed.2006.06.042

33. Jackson CA, Sudlow CLM, Mishra GD. Education, sex and risk of stroke: a prospective cohort study in New South Wales, Australia. BMJ Open. 2018;8:e024070. 10.1136/bmjopen-2018-024070

34. Xu N, Qiu Y, Ainiwan D, Wang B, Alifu X, Zhou H, et al. Mediating factors in the association between educational attainment and stroke: A mendelian randomization study. SSM - Popul Health. 2025;29:101766. 10.1016/j.ssmph.2025.101766

35. Teppo K, Jaakkola J, Biancari F, Halminen O, Linna M, Haukka J, et al. Association of income and educational levels with adherence to direct oral anticoagulant therapy in patients with incident atrial fibrillation: A Finnish nationwide cohort study. Pharmacol Res Perspect. 2022;10:e00961. 10.1002/prp2.961

36. Humphries KH, Kerr CR, Connolly SJ, Klein G, Boone JA, Green M, et al. New-Onset Atrial Fibrillation.

37. Dagres N, Nieuwlaat R, Vardas PE, Andresen D, Lévy S, Cobbe S, et al. Gender-Related Differences in Presentation, Treatment, and Outcome of Patients With Atrial Fibrillation in Europe. J Am Coll Cardiol. 2007;49:572–7. 10.1016/j.jacc.2006.10.047

38. Emdin CA, Wong CX, Hsiao AJ, Altman DG, Peters SA, Woodward M, et al. Atrial fibrillation as risk factor for cardiovascular disease and death in women compared with men: systematic review and meta-analysis of cohort studies. BMJ. 2016;h7013. 10.1136/bmj.h7013

